# Effectiveness of primary-contact physiotherapy in managing musculoskeletal conditions in emergency departments: Statistical analysis plan for the RESHAP-ED randomised controlled trial

**DOI:** 10.1101/2025.11.06.25339640

**Authors:** Sana Shan, Laurent Billot, Gustavo Machado, Chris Maher

## Abstract

The RESHAP-ED^1^ trial is a pragmatic, multicentre, two-arm, parallel randomised controlled trial which aims to test the effectiveness of primary-contact physiotherapy for musculoskeletal pain when compared to the traditional doctor/nurse pathway, on patient flow (i.e. length of stay) in the emergency department. The objective of this trial is to compare two approaches to physiotherapy involvement in the assessment and management of patients who present to emergency departments with simple musculoskeletal problems that are both standard of care in Australian emergency departments.

The primary health services outcome is emergency department length of stay defined as the period between when a patient presents at an emergency department, and when that person is recorded as having physically departed the emergency department. The primary outcome will be analysed using a generalised linear mixed effects model with a fixed effect of intervention (primary-contact physiotherapy vs control) and a random effect for hospital. The statistical analysis plan pre-specifies the method of analysis for primary, secondary and safety outcomes and key variables collected in the trial.

Secondary, and subgroup group analyses have been pre-specified as well.

## 1 Administrative information

### 1.1 Study identifiers

Australian New Zealand Clinical Trials Registry. Identifier: ACTRN12623000782639, Date: 18/07/2023

### 1.2 Revision history

| Version | Date | Details |
| --- | --- | --- |
| 0.1 (draft) | 21 May 2025 | First draft by Sana |
| 1.0 | 28 August 2025 | Second draft by Sana |
| Final | 10 October 2025 | Final draft by Sana |

### 1.3 Contributors to the statistical analysis plan

#### 1.3.1 Roles and responsibilities

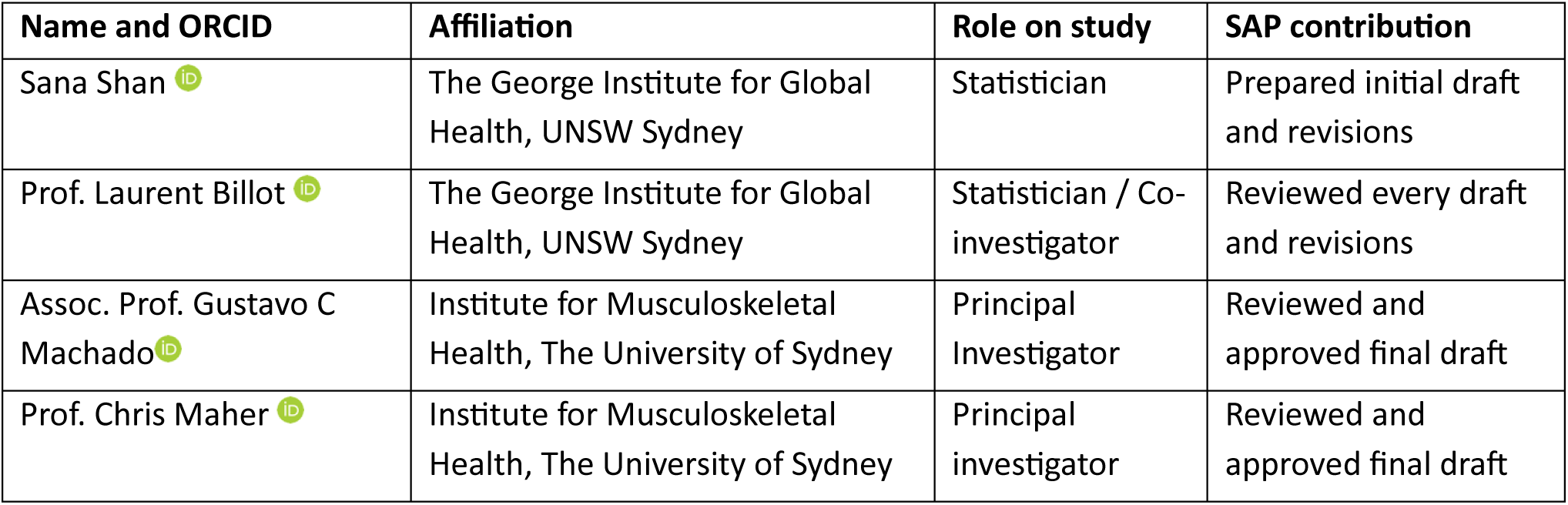

1.3 Contributors to the statistical analysis plan 1.3.1 Roles and responsibilities
| Name and ORCID | Affiliation | Role on study | SAP contribution |
| --- | --- | --- | --- |
| Sana Shan | The George Institute for Global Health, UNSW Sydney | Statistician | Prepared initial draft and revisions |
| Prof. Laurent Billot | The George Institute for Global Health, UNSW Sydney | Statistician / Co-investigator | Reviewed every draft and revisions |
| Assoc. Prof. Gustavo C Machado | Institute for Musculoskeletal Health, The University of Sydney | Principal Investigator | Reviewed and approved final draft |
| Prof. Chris Maher | Institute for Musculoskeletal Health, The University of Sydney | Principal investigator | Reviewed and approved final draft |

#### 1.3.2 Approvals

The undersigned have reviewed this plan and approve it as final. They find it to be consistent with the requirements of the protocol as it applies to their respective areas. They also find it to be compliant with ICH-E9 principles and in particular, confirm that this analysis plan was developed in a completely blinded manner, i.e. without knowledge of the effect of the intervention(s) being assessed.

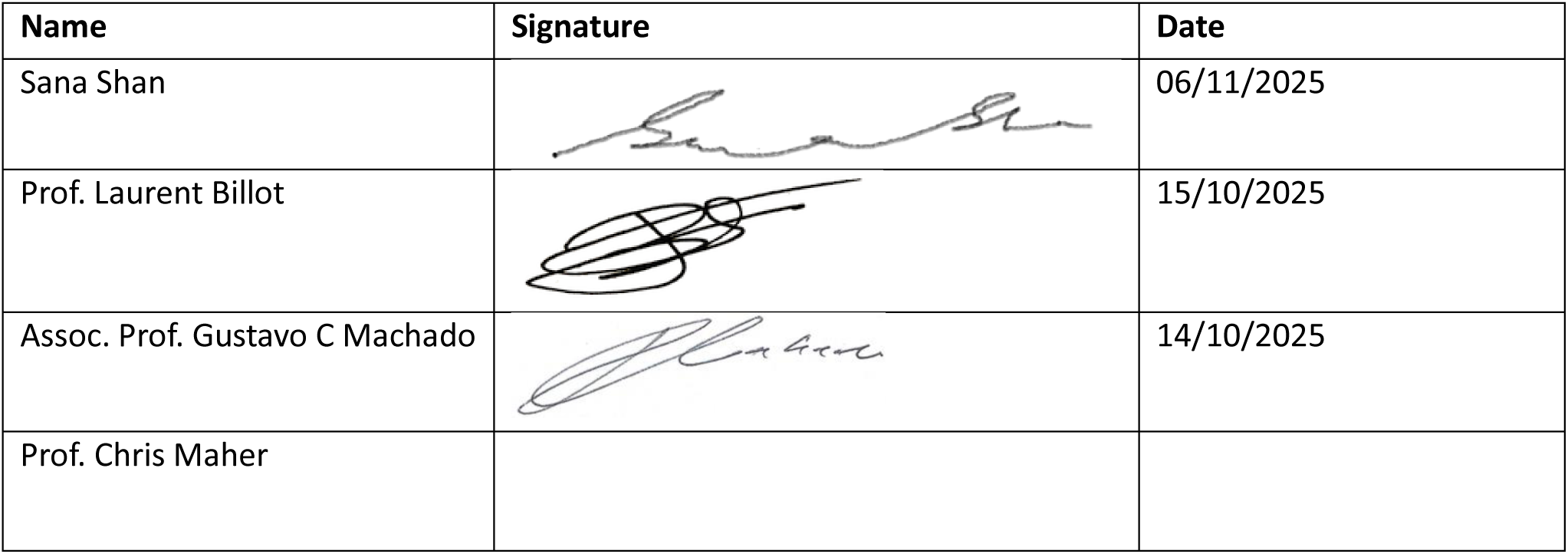

| Name | Signature | Date |
| --- | --- | --- |
| Sana Shan |  | 06/11/2025 |
| Prof. Laurent Billot |  | 15/10/2025 |
| Assoc. Prof. Gustavo C Machado |  | 14/10/2025 |
| Prof. Chris Maher |  |  |

## 2 Introduction

### 2.2 Study population

This trial will be conducted in multiple NSW public hospital emergency departments from Sydney metropolitan, and regional and rural areas of NSW. These emergency departments provide diversity with respect to patient demographic characteristics, ethnicity, hospital setting, and emergency department clinical staffing models. We expect to include 1370 participants in this trial across 5 NSW public hospitals.

#### 2.2.1 Inclusion criteria

**Hospital level**

These are the eligibility criteria at a hospital level:

- Departmental agreement to implement a primary-contact physiotherapy pathway and the appropriate governance if they do not already have this service; and
- An electronic medical record system.

**Individual level**

***Inclusion criteria***

- Adult patient (≥18 years);
- Presents to the emergency department with a simple musculoskeletal condition. This would include, but is not limited to, soft tissue injuries (joint, ligament, tendon or muscle pain), musculoskeletal neck and low back pain, closed peripheral fractures not requiring reduction or orthopaedic fixation (indicated from triage notes), spontaneously reduced joint dislocations; and
- Triage score 3 (urgent), 4 (semi-urgent) or 5 (non-urgent) as per Australian Triage Scale. Triage score 2 (emergency) will also be included whenever an adult presents to the emergency department with an isolated joint dislocation including: shoulder, digit, patella or temporomandibular joint (indicated by triage notes).

***Exclusion criteria***

- Already assessed by a doctor or other primary-contact health practitioner in the emergency department as indicated on the Cerner FirstNet® list; or
- Any non-musculoskeletal/non-orthopaedic condition, such as open wounds, eye problems, foreign bodies, and poisonings; or
- Presentations outside the primary-contact physiotherapists’ working hours; or
- Triage notes indication of soft tissue injuries that may require surgical intervention; or
- Triage notes indication of fracture requiring reduction and/or surgical fixation; or
- Triage notes indication of signs or symptoms of serious pathology (for example: spinal fracture, cauda equina syndrome, malignancy, infection, inflammatory origin of pain); or
- Co-morbidities requiring immediate emergency care (for example: cardiac, respiratory; or psychiatric issues as indicated on the triage note); or
- Unable to provide sufficient informed consent to participate (eg. cognitive impairment, intoxication, unable to understand spoken or written English).
- Patient re-presentation that has already been included in the trial.

## 3 Study interventions

The primary-contact physiotherapy intervention involves assessment and management of musculoskeletal cases by a primary-contact senior level 4 physiotherapist who will assess and manage the patient as primary contact. After their initial assessment, the physiotherapist will choose the best course of management with consultation with the patient and carer. Care for the low back pain cases will be provided according to recommendations from the Australian Low Back Pain Clinical Care Standard^2^, and other musculoskeletal conditions will be managed via best practice as recommended by the Emergency Care Institute^3^. The physiotherapist may consult a medical/nurse practitioner (collaborative model of care) or autonomously manage eligible patients. In the autonomous model of care, the physiotherapist is responsible for selecting eligible patients from the triage list, assessment, management, diagnosis, and discharge with no involvement of a medical/nurse practitioner. In the collaborative model of care, after selecting and assessing the patient, the primary-contact physiotherapist shares discharge and management decision making with a medical/nurse practitioner.

The control group consists of primary-contact care pathway by a doctor and/or nurse practitioner. The care provided will be decided by the emergency department doctors and nurses, as per usual. Emergency doctors and/or nurses may refer the patients to see the physiotherapist in the emergency department as secondary-contact, or to the outpatient physiotherapy department after discharge.

### 3.1 Randomisation

The randomisation unit for intervention or control groups is the patient. Computer-generated random numbers will be used to assign eligible participants to the intervention or control group in a 1:1 ratio. Randomisation will be performed online by the primary-contact physiotherapist.

## 4 Outcomes

### 4.1 Primary outcome

The primary health services outcome is emergency department length of stay defined as the period between when a patient presents at an emergency department, and when that person is recorded as having physically departed the emergency department.

### 4.2 Secondary outcomes

#### 4.2.1 Health services outcomes

- Time to clinical care (i.e. time elapsed in minutes for each patient from presentation in the emergency department to the commencement of the emergency department non-admitted clinical care. METeOR id: 621840).
- Duration of clinical care (i.e. the period between when clinical care commences and the end of the non-admitted patient emergency department episode, i.e. physical departure from the emergency department).
- Proportion of patients whose emergency department stay (as defined above) is less than or equal to four hours (proportion completed within four hours).
- Proportion of patients ‘seen on time’ (i.e. the proportion of presentations for which the waiting time to commencement of clinical care was within the time specified in the definition of the triage category: emergency (within 10 minutes), urgent (within 30 minutes), semi-urgent (within 60 minutes), non-urgent (within 120 minutes)).
- Proportion of patients admitted to EDSSU (Emergency Department Short Stay Unit).
- Proportion of patients admitted to hospital inpatient units.
- Proportion of patients that received imaging.
- Proportion of patients that received opioids.
- Proportion of patients that received gabapentinoids.
- Proportion of patients that received benzodiazepines.
- Proportion of patients re-presenting to the emergency department for the same chief complaint within 72 hours of discharge.
- Proportion of patients readmitted to hospital inpatient units for the same chief complaint within 30 days of index inpatient admission.

#### 4.2.2 Self-reported outcomes

- Pain intensity (numeric rating scale, range 0–10).
- Quality of life (Euro-Qol-5D).
- Satisfaction with care (1 item from the Emergency Department Patient Experience of Care (EDPEC) Survey).

### 4.3 Safety outcomes

- Subsequent hospitalisations from the emergency department.
- Unplanned re-presentation to the emergency department related to the trial presentation within 72 hours and 6 weeks. For example, due to missed fracture diagnosis or ongoing pain.
- Adverse events reported on day 1 after emergency department discharge.

## 5 Analysis principles

### 5.1 Sample size

Initially, it was estimated that with a total sample size of 620 patients (310 per arm), the study will have 90% power to detect a difference of 30 minutes in emergency department length of stay assuming a standard-deviation of 115 minutes. A re-estimation of sample size was performed based on reassessment of diagnosis codes to account for patient presentations with fracture/dislocation. Based on this re-estimation, a sample size of 1370 will have 90% power overall to detect a 30-minute difference based on the SD of 125 minutes (based on the observed SD from initial descriptive analysis of the study). We will have at least 80% power within each of the two main subgroups: those with fracture/dislocation (n=548) vs soft tissue injuries/other (n=822) enabling reasonably well-powered subgroup analyses by hospital.

### 5.2 Software

Analyses will be conducted primarily using SAS Enterprise Guide (version 8.3 or above) and R (version 4.0.0 or above).

### 5.3 Interim analyses

No formal interim analyses were conducted during the study.

### 5.4 Multiplicity adjustment

Statistical tests are to be two-sided with a nominal level of *α* set at 5%. Analysis of the primary outcome will be unadjusted for multiplicity. For the 12 secondary health services outcomes measured, we will control the family-wise error rate by grouping health services endpoints by family (e.g. ED flow family, disposition and resource use family and analgesia and imaging family) and apply a sequential Holm-Sidak correction^4^. All p-values will be ranked from smallest to largest and then comparing them to an adjusted level of significance calculated as 1-(1-0.05),1/C where C indicates the number of comparisons that remain. The sequential testing procedure stops as soon as a p value fails to reach the corrected significance level. All other secondary endpoints including self-reported outcomes will be considered exploratory in nature and will not be adjusted for multiplicity.

Secondary endpoint families will be defined as follows:

**Family 1: ED Flow**

- Time to clinical care
- Duration of clinical care
- Proportion left ≤ 4 hours
- Proportion ‘seen on time’

**Family 2: ED Disposition and Resource Use**

- Proportion admitted to EDSSU
- Proportion admitted to inpatient units
- Proportion re-presenting to the ED within 72 hours
- Proportion readmitted to hospital within 30 days

**Family 3: Analgesia and Imaging**

- Proportion that received imaging
- Proportion that received opioids
- Proportion that received gabapentinoids
- Proportion that received benzodiazepines

### 5.5 Datasets analysed

#### 5.5.1 Analysis population

The intention-to-treat (ITT) analysis set will be used to assess both effectiveness and safety.

#### 5.5.2 Analysis strategy

For all outcomes, the analyses will be performed on the ITT population using all available data. A waiver of consent will be requested to collect routinely collected data from electronic medical records to describe the characteristics of the patients and episodes of care and to collect the health services outcomes. This will be extracted retrospectively after the patient leaves the emergency departments. This will also ensure minimum missing data for the primary outcome; therefore, no imputation is planned for missing data.

## 6 Planned Analyses

### 6.1 Subject disposition

The flow of patients through the trial will be displayed in a CONSORT^5^ (Consolidated Standards of Reporting Trials) diagram [see Figure 1]. The report will include the following: the number of participants assessed for eligibility, reasons for exclusion, the number of enrolled participants, the number and reasons for post-randomisation exclusions, and the proportion of eligible patients being enrolled in each treatment group of the study. The flow chart will also include the number of patients alive and available at end of follow-up (Day 42).

### 6.2 Baseline comparison

#### 6.2.1 Hospital characteristics

Description of the hospital characteristics (e.g. no. of beds (above and below median values), hospital location (regional, metropolitan), and annual number presentations for all conditions to emergency departments (above and below median values)) will be presented. Discrete variables will be summarised by frequencies and percentages. Percentages will be calculated according to the number of clusters with available data. Continuous variables will be summarised by using mean and standard deviation (SD), and median and interquartile range (Q1-Q3).

#### 6.2.2 Patient characteristics

Description of baseline patient characteristics will be presented by treatment group. Discrete variables will be summarised by frequencies and percentages. Percentages will be calculated according to the number of patients in whom data are available. Continuous variables will be summarised by using mean and SD, and median and interquartile range (Q1-Q3). No adjustment for clustering will be applied when summarising baseline characteristics. Baseline measures for all patients will be tabulated for the variables listed below

Baseline measures for all eligible musculoskeletal pain presentations will be tabulated with at least the following variables outlined below:

- Age
- Sex
- ED Diagnosis
- Day of ED presentation
- ED presentation hour (Normal (not ED) hours vs. ED hours)
- ED mode of arrival
- ED triage category

### 6.3 Follow-up assessments

All assessments/ online surveys carried out during follow-up will be described by treatment group. Details are presented in mock tables. No formal statistical tests are planned for these variables.

### 6.4 Analysis of the primary outcome

***Main analysis***

To analyse the primary outcome of emergency department length of stay, a continuous variable will be created indicating the time in hours from when a patient presents at an emergency department and when that person is recorded as having physically departed the emergency department. The intervention effect will be estimated using a generalised linear mixed effects model using a gaussian distribution and an identity link function. The model will include a fixed effect of intervention (primary-contact physiotherapy vs control) and a random effect for hospital. The results will be presented as mean difference (MD) between the treatment groups with 95% confidence intervals (CI).

### 6.5 Adjusted analyses

Adjusted analyses of the primary outcome will be performed by adding the following baseline covariates to the main analysis model (see section 6.4): age, sex, type of musculoskeletal injury (soft tissue vs. fracture). The adjusted treatment effect will be reported as the mean difference (MD) and 95% CI.

### 6.6 Sensitivity analyses

While length of stay is unlikely to follow a normal distribution, the large sample size will allow us to make inference about the mean. In case of important heteroscedasticity, as assessed using a plot of residuals against fitted values, we will conduct a sensitivity analysis using a log-transformation.

### 6.7 Treatment of missing data

We are not planning to conduct any multiple imputation as these data will be extracted from the electronic medical record system and no or very little (<5%) missingness is expected for the primary outcomes of emergency department length of stay.

### 6.8 Per-protocol analysis

No per-protocol analysis will be carried out.

### 6.9 Analysis of the secondary outcomes

Analysis of secondary health services continuous outcomes will follow the same analysis approach as the one used for the primary outcome main analysis, see section 6.4 for details. For binary outcomes, gaussian distribution will be replaced by binary distribution and logit link function. Results of binary outcomes will be presented as odds ratios and 95% confidence intervals. No adjustment of covariates is planned for secondary outcomes.

Secondary self-reported health outcomes (including EQ5D domains and health utility index, pain intensity and satisfaction of care) will be analysed using a generalised linear mixed effects model with gaussian distribution and an identity link function. The model will include a fixed effect of intervention (primary-contact physiotherapy vs control), fixed effect of time as a categorical variable (Day 1, Day 7 and Day 42), a fixed treatment and time interaction and a random effect for hospital. Using contrasts at specific timepoints, the results will be presented as mean difference (MD) between the treatment groups with 95% confidence intervals (CI).

### 6.10 Analysis by musculoskeletal injury diagnosis

Randomised patients will be split into two cohorts: those who presented to the emergency department with a soft tissue / other injury and those who presented with a fracture/dislocation. Adjudication of the presenting musculoskeletal condition will be done by the study team. Analysis of the primary and health services secondary outcomes will be repeated separately within each cohort.

### 6.11 Analysis of safety outcomes

Safety outcomes of all-cause and cause-specific SAEs will be summarised as the number and proportion of patients experiencing at least one event. Adverse events will be summarised by category of event and overall numbers of events. In addition to the number of patients with at least one event, we will report the total number of events.

A listing of all AEs and SAEs will be reported (in an appendix).

### 6.12 Subgroup analyses

Heterogeneity of treatment on the primary endpoint will be assessed in the following pre-defined subgroups:

- age category (<65 vs. ≥65 years)
- sex
- hospital
- musculoskeletal injury (fractures/dislocation vs. soft tissue injury)

The analysis for each subgroup will be performed by adding the subgroup variable as well as its interaction with the intervention as fixed effects to the main analysis model for primary outcome, as described in section 6.4. Within each subgroup, summary measures will include frequency of events and percentage, within each treatment arm, as well as the mean difference (MD) for treatment effect with a 95% CI. The results will be displayed on a forest plot [figure 2] including the p-value for heterogeneity corresponding to the interaction term between the intervention and the subgroup variable.

## Data Availability

N/A

## 8 Appendix 1 Proposed main tables and figures

**Figure 1:**
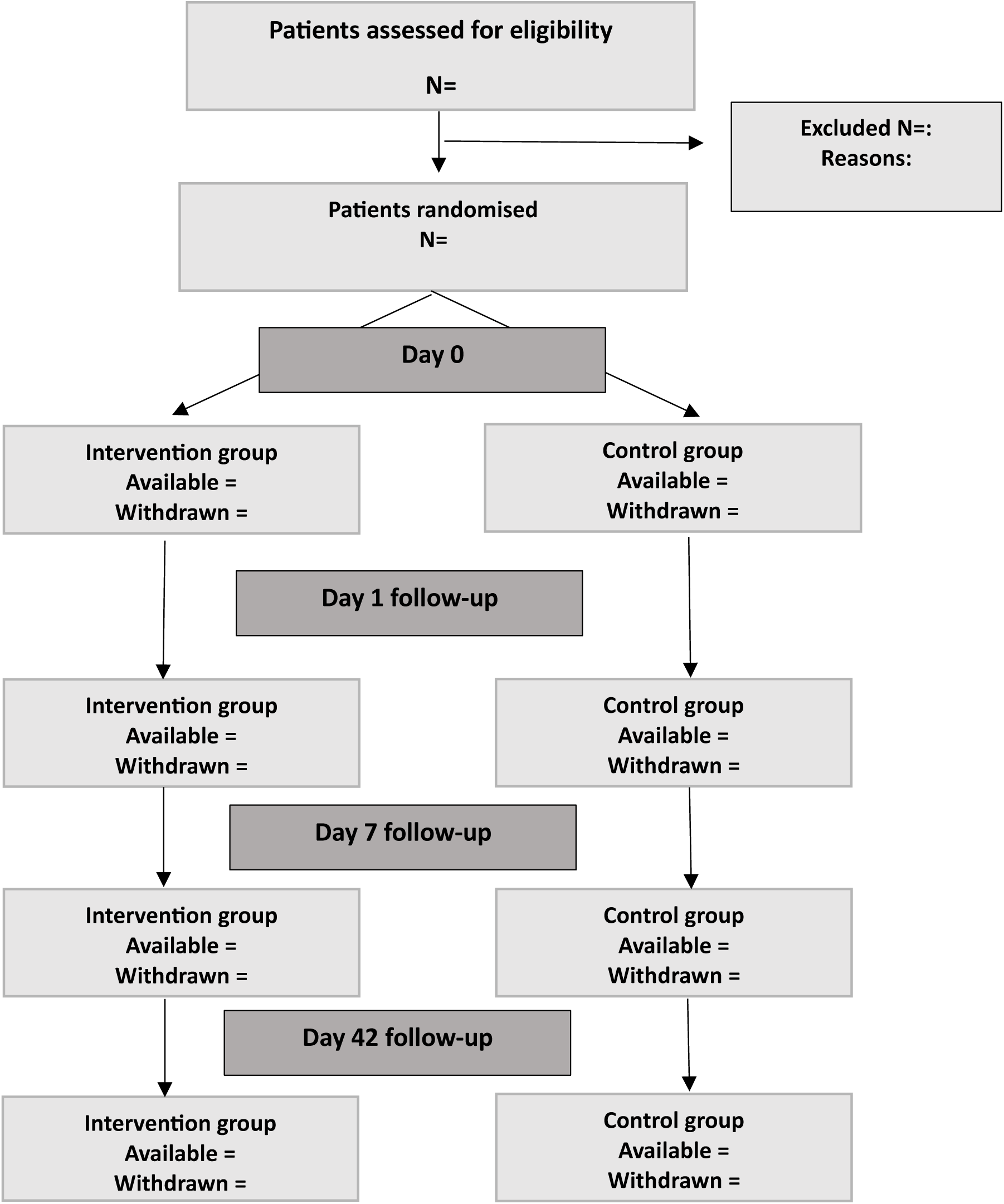
Consort flowchart.

**Table 1:** Hospital characteristics.

| Hospital characteristics | Total |
| --- | --- |
| **Location** | N=xxx |
| Metropolitan | xx (xx.x%) |
| Regional | xx (xx.x%) |
| **No. hospital beds** |  |
| N Mean(SD) | N xx.x (xx.x) |
| Median [Q1 – Q3] | xx.x [xx.x- xx.x] |
| **No. of beds above median** | xx (xx.x%) |
| **No. of beds below median** | xx (xx.x%) |
| **Annual ED presentations for all conditions** |  |
| N Mean (SD) | N xx.x (xx.x) |
| Median [Q1 – Q3] | xx.x [xx.x- xx.x] |
| **ED presentations above median** | xx (xx.x%) |
| **ED presentations below median** | xx (xx.x%) |

**Table 2:** Subject disposition.

|  | Intervention | Control | Total |
| --- | --- | --- | --- |
| **Randomised patients** | N=xx | N=xx | N=xx |
| Withdrawn from trial at day 0 | xx (xx.x%) | xx (xx.x%) | xx (xx.x%) |
| Withdrawn from trial day 1 | xx (xx.x%) | xx (xx.x%) | xx (xx.x%) |
| Withdrawn from trial day 7 | xx (xx.x%) | xx (xx.x%) | xx (xx.x%) |
| Withdrawn from trial at day 42 | xx (xx.x%) | xx (xx.x%) | xx (xx.x%) |
| **Post randomisation exclusion** | xx (xx.x%) | xx (xx.x%) | xx (xx.x%) |
| Reasons for exclusion |  |  |  |
| Reason 1 | xx (xx.x%) | xx (xx.x%) | xx (xx.x%) |
| Reason 2 | xx (xx.x%) | xx (xx.x%) | xx (xx.x%) |
| **Completed the study** |  |  |  |
| Yes | xx (xx.x%) | xx (xx.x%) | xx (xx.x%) |
| No | xx (xx.x%) | xx (xx.x%) | xx (xx.x%) |
| Partial | xx (xx.x%) | xx (xx.x%) | xx (xx.x%) |
| **Reasons for withdrawal** | N=xx | N=xx | N=xx |
| xxx | xx (xx.x%) | xx (xx.x%) | xx (xx.x%) |
| xxx | xx (xx.x%) | xx (xx.x%) | xx (xx.x%) |
| Other | xx (xx.x%) | xx (xx.x%) | xx (xx.x%) |

**Table 3:** Baseline characteristics table.

| Characteristics | Intervention | Control | Total |
| --- | --- | --- | --- |
| **Age (years)** |  |  |  |
| N Mean (SD) | N xx.x (xx.x) | N xx.x (xx.x) | N xx.x (xx.x) |
| Median [Q1 – Q3] | xx.x [xx.x- xx.x] | xx.x [xx.x- xx.x] | xx.x [xx.x- xx.x] |
| **Sex** | N=xxx | N=xxx | N=xxx |
| Male | xx (xx.x%) | xx (xx.x%) | xx (xx.x%) |
| Female | xx (xx.x%) | xx (xx.x%) | xx (xx.x%) |
| **Country of birth** | N=xxx | N=xxx | N=xxx |
| Australia | xx (xx.x%) | xx (xx.x%) | xx (xx.x%) |
| Overseas | xx (xx.x%) | xx (xx.x%) | xx (xx.x%) |
| **Language spoken at home** |  |  |  |
| English | xx (xx.x%) | xx (xx.x%) | xx (xx.x%) |
| Non-english | xx (xx.x%) | xx (xx.x%) | xx (xx.x%) |
| **Need for interpreter** | xx (xx.x%) | xx (xx.x%) | xx (xx.x%) |
| **Postcode of residence** |  |  |  |
| SEIFA category (1-5) | xx (xx.x%) | xx (xx.x%) | xx (xx.x%) |
| SEIFA category (6-10) | xx (xx.x%) | xx (xx.x%) | xx (xx.x%) |

**Table 4:**
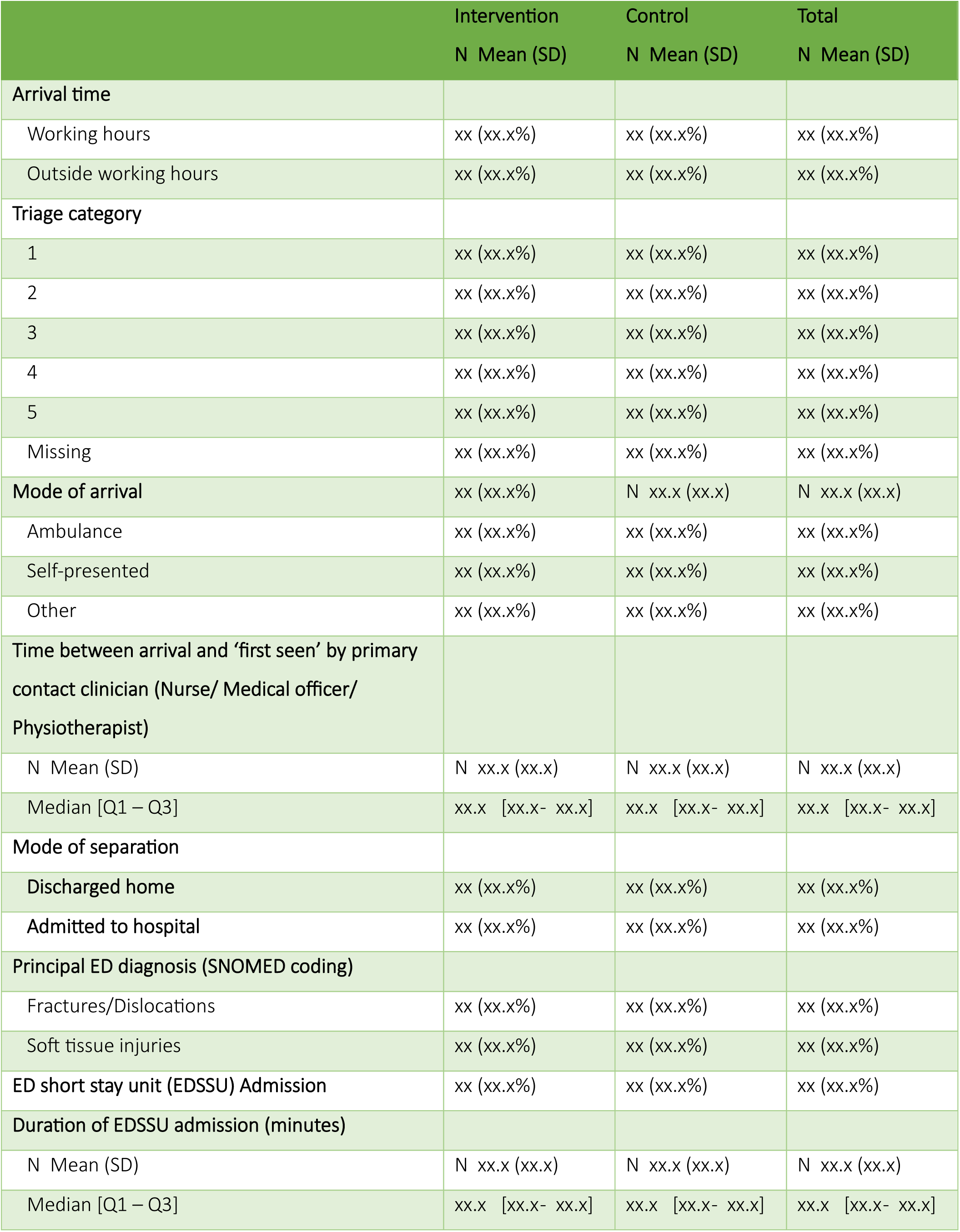
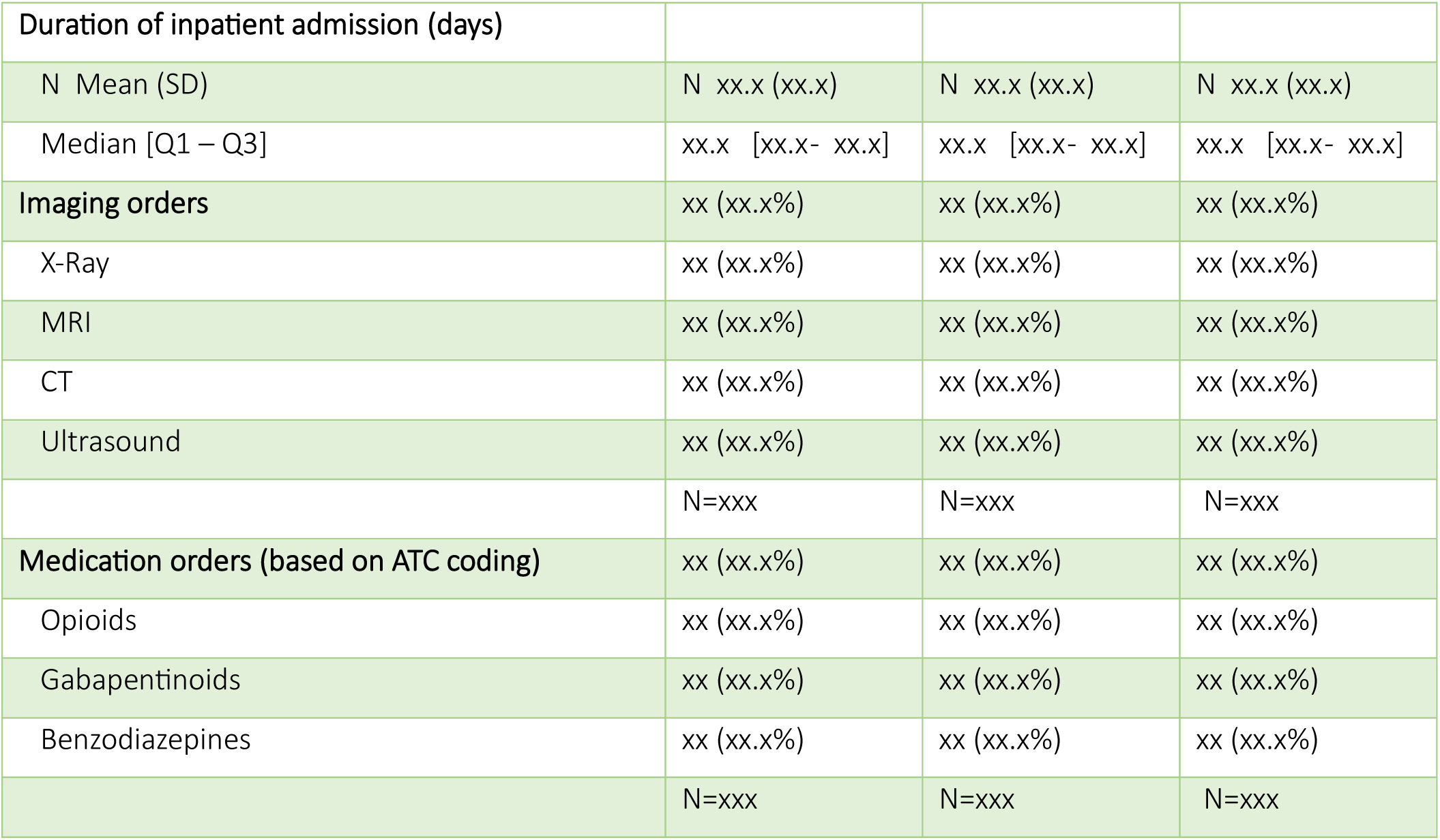
Summary of Electronic Medical Record data.

|  | Intervention | Control | Total |
| --- | --- | --- | --- |
|  | N Mean (SD) | N Mean (SD) | N Mean (SD) |
| **Arrival time** |  |  |  |
| Working hours | xx (xx.x%) | xx (xx.x%) | xx (xx.x%) |
| Outside working hours | xx (xx.x%) | xx (xx.x%) | xx (xx.x%) |
| **Triage category** |  |  |  |
| 1 | xx (xx.x%) | xx (xx.x%) | xx (xx.x%) |
| 2 | xx (xx.x%) | xx (xx.x%) | xx (xx.x%) |
| 3 | xx (xx.x%) | xx (xx.x%) | xx (xx.x%) |
| 4 | xx (xx.x%) | xx (xx.x%) | xx (xx.x%) |
| 5 | xx (xx.x%) | xx (xx.x%) | xx (xx.x%) |
| Missing | xx (xx.x%) | xx (xx.x%) | xx (xx.x%) |
| **Mode of arrival** | xx (xx.x%) | N xx.x (xx.x) | N xx.x (xx.x) |
| Ambulance | xx (xx.x%) | xx (xx.x%) | xx (xx.x%) |
| Self-presented | xx (xx.x%) | xx (xx.x%) | xx (xx.x%) |
| Other | xx (xx.x%) | xx (xx.x%) | xx (xx.x%) |
| **Time between arrival and ‘first seen’ by primary contact clinician (Nurse/ Medical officer/ Physiotherapist)** |  |  |  |
| N Mean (SD) | N xx.x (xx.x) | N xx.x (xx.x) | N xx.x (xx.x) |
| Median [Q1 – Q3] | xx.x [xx.x- xx.x] | xx.x [xx.x- xx.x] | xx.x [xx.x- xx.x] |
| **Mode of separation** |  |  |  |
| Discharged home | xx (xx.x%) | xx (xx.x%) | xx (xx.x%) |
| Admitted to hospital | xx (xx.x%) | xx (xx.x%) | xx (xx.x%) |
| **Principal ED diagnosis (SNOMED coding)** |  |  |  |
| Fractures/Dislocations | xx (xx.x%) | xx (xx.x%) | xx (xx.x%) |
| Soft tissue injuries | xx (xx.x%) | xx (xx.x%) | xx (xx.x%) |
| **ED short stay unit (EDSSU) Admission** | xx (xx.x%) | xx (xx.x%) | xx (xx.x%) |
| **Duration of EDSSU admission (minutes)** |  |  |  |
| N Mean (SD) | N xx.x (xx.x) | N xx.x (xx.x) | N xx.x (xx.x) |
| Median [Q1 – Q3] | xx.x [xx.x- xx.x] | xx.x [xx.x- xx.x] | xx.x [xx.x- xx.x] |
| **Duration of inpatient admission (days)** |  |  |  |
| N Mean (SD) | N xx.x (xx.x) | N xx.x (xx.x) | N xx.x (xx.x) |
| Median [Q1 – Q3] | xx.x [xx.x- xx.x] | xx.x [xx.x- xx.x] | xx.x [xx.x- xx.x] |
| **Imaging orders** | xx (xx.x%) | xx (xx.x%) | xx (xx.x%) |
| X-Ray | xx (xx.x%) | xx (xx.x%) | xx (xx.x%) |
| MRI | xx (xx.x%) | xx (xx.x%) | xx (xx.x%) |
| CT | xx (xx.x%) | xx (xx.x%) | xx (xx.x%) |
| Ultrasound | xx (xx.x%) | xx (xx.x%) | xx (xx.x%) |
|  | N=xxx | N=xxx | N=xxx |
| **Medication orders (based on ATC coding)** | xx (xx.x%) | xx (xx.x%) | xx (xx.x%) |
| Opioids | xx (xx.x%) | xx (xx.x%) | xx (xx.x%) |
| Gabapentinoids | xx (xx.x%) | xx (xx.x%) | xx (xx.x%) |
| Benzodiazepines | xx (xx.x%) | xx (xx.x%) | xx (xx.x%) |
|  | N=xxx | N=xxx | N=xxx |

**Table 5:** Patient reported outcomes – descriptives by timepoints.

|  | Intervention | Control | Total |
| --- | --- | --- | --- |
| **Day 1** |  |  |  |
| **EQ5d-5L** | N=xxx | N=xxx | N=xxx |
| **Self-reported general health VAS (100=best health)** |  |  |  |
| N Mean (SD) | N xx.x(xx.x) | N xx.x (xx.x) | N xx.x (xx.x) |
| Median [Q1 – Q3] | xx.x [xx.x- xx.x] | xx.x [xx.x- xx.x] | xx.x [xx.x- xx.x] |
| **Health utility score** |  |  |  |
| N Mean (SD) | N xx.x(xx.x) | N xx.x (xx.x) | N xx.x (xx.x) |
| Median [Q1 – Q3] | xx.x [xx.x- xx.x] | xx.x [xx.x- xx.x] | xx.x [xx.x- xx.x] |
| **Pain intensity** | N=xxx | N=xxx | N=xxx |
| N Mean (SD) | N xx.x(xx.x) | N xx.x (xx.x) | N xx.x (xx.x) |
| Median [Q1 – Q3] | xx.x [xx.x- xx.x] | xx.x [xx.x- xx.x] | xx.x [xx.x- xx.x] |
| **Satisfaction with care** | N=xxx | N=xxx | N=xxx |
| N Mean (SD) | N xx.x(xx.x) | N xx.x (xx.x) | N xx.x (xx.x) |
| Median [Q1 – Q3] | xx.x [xx.x- xx.x] | xx.x [xx.x- xx.x] | xx.x [xx.x- xx.x] |
| **Day 7** |  |  |  |
| **EQ5d-5L** | N=xxx | N=xxx | N=xxx |
| **Self-reported general health VAS (100=best health)** |  |  |  |
| N Mean (SD) | N xx.x(xx.x) | N xx.x (xx.x) | N xx.x (xx.x) |
| Median [Q1 – Q3] | xx.x [xx.x- xx.x] | xx.x [xx.x- xx.x] | xx.x [xx.x- xx.x] |
| **Health utility score** |  |  |  |
| N Mean (SD) | N xx.x(xx.x) | N xx.x (xx.x) | N xx.x (xx.x) |
| Median [Q1 – Q3] | xx.x [xx.x- xx.x] | xx.x [xx.x- xx.x] | xx.x [xx.x- xx.x] |
| **Pain intensity** | N=xxx | N=xxx | N=xxx |
| N Mean (SD) | N xx.x(xx.x) | N xx.x (xx.x) | N xx.x (xx.x) |
| Median [Q1 – Q3] | xx.x [xx.x- xx.x] | xx.x [xx.x- xx.x] | xx.x [xx.x- xx.x] |
| **Satisfaction with care** | N=xxx | N=xxx | N=xxx |
| N Mean (SD) | N xx.x(xx.x) | N xx.x (xx.x) | N xx.x (xx.x) |
| Median [Q1 – Q3] | xx.x [xx.x- xx.x] | xx.x [xx.x- xx.x] | xx.x [xx.x- xx.x] |

| Day 42 |  |  |  |
| --- | --- | --- | --- |
| **EQ5d-5L** | N=xxx | N=xxx | N=xxx |
| **Self-reported general health VAS (100=best health)** |  |  |  |
| N Mean (SD) | N xx.x(xx.x) | N xx.x (xx.x) | N xx.x (xx.x) |
| Median [Q1 – Q3] | xx.x [xx.x- xx.x] | xx.x [xx.x- xx.x] | xx.x [xx.x- xx.x] |
| **Health utility score** |  |  |  |
| N Mean (SD) | N xx.x(xx.x) | N xx.x (xx.x) | N xx.x (xx.x) |
| Median [Q1 – Q3] | xx.x [xx.x- xx.x] | xx.x [xx.x- xx.x] | xx.x [xx.x- xx.x] |
| **Pain intensity** | N=xxx | N=xxx | N=xxx |
| N Mean (SD) | N xx.x(xx.x) | N xx.x (xx.x) | N xx.x (xx.x) |
| Median [Q1 – Q3] | xx.x [xx.x- xx.x] | xx.x [xx.x- xx.x] | xx.x [xx.x- xx.x] |
| **Satisfaction with care** | N=xxx | N=xxx | N=xxx |
| N Mean (SD) | N xx.x(xx.x) | N xx.x (xx.x) | N xx.x (xx.x) |
| Median [Q1 – Q3] | xx.x [xx.x- xx.x] | xx.x [xx.x- xx.x] | xx.x [xx.x- xx.x] |

**Table 6:**
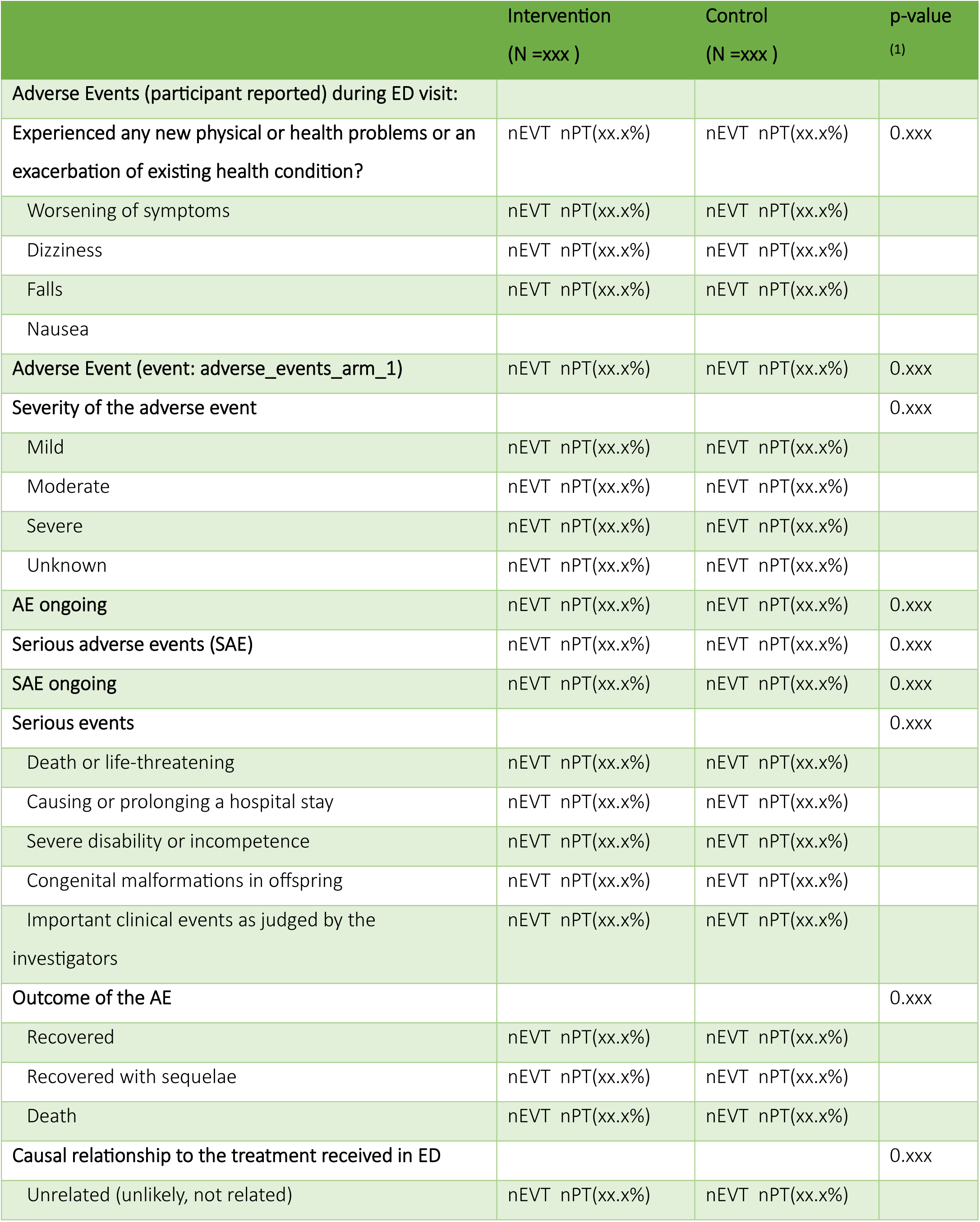
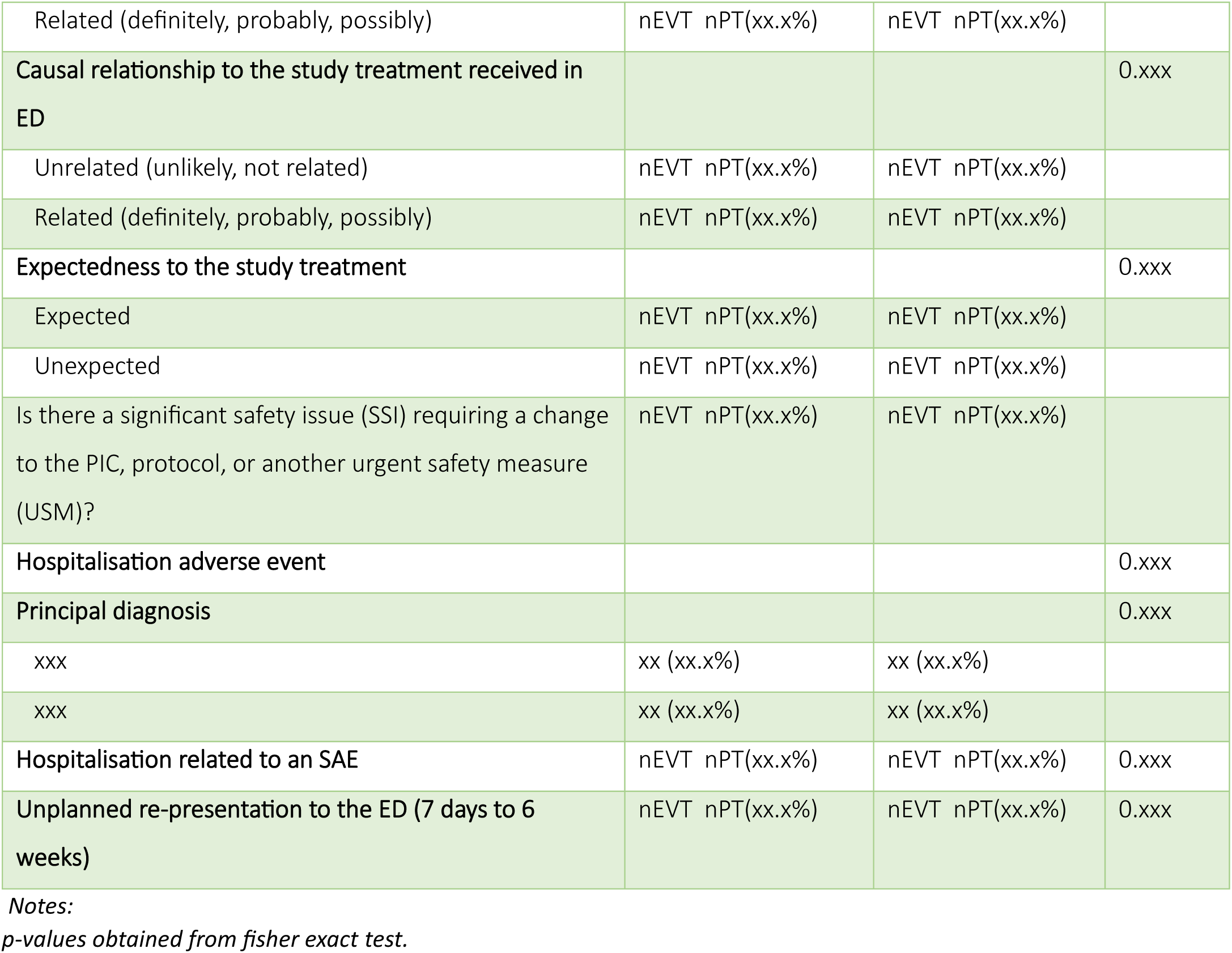
Summary of adjudicated adverse events.

|  | Intervention<br>(N =xxx ) | Control<br>(N =xxx ) | p-value<br>(1) |
| --- | --- | --- | --- |
| **Adverse Events (participant reported) during ED visit:** |  |  |  |
| **Experienced any new physical or health problems or an exacerbation of existing health condition?** | nEVT nPT(xx.x%) | nEVT nPT(xx.x%) | 0.xxx |
| Worsening of symptoms | nEVT nPT(xx.x%) | nEVT nPT(xx.x%) |  |
| Dizziness | nEVT nPT(xx.x%) | nEVT nPT(xx.x%) |  |
| Falls | nEVT nPT(xx.x%) | nEVT nPT(xx.x%) |  |
| Nausea |  |  |  |
| **Adverse Event (event: adverse_events_arm_1)** | nEVT nPT(xx.x%) | nEVT nPT(xx.x%) | 0.xxx |
| **Severity of the adverse event** |  |  | 0.xxx |
| Mild | nEVT nPT(xx.x%) | nEVT nPT(xx.x%) |  |
| Moderate | nEVT nPT(xx.x%) | nEVT nPT(xx.x%) |  |
| Severe | nEVT nPT(xx.x%) | nEVT nPT(xx.x%) |  |
| Unknown | nEVT nPT(xx.x%) | nEVT nPT(xx.x%) |  |
| **AE ongoing** | nEVT nPT(xx.x%) | nEVT nPT(xx.x%) | 0.xxx |
| **Serious adverse events (SAE)** | nEVT nPT(xx.x%) | nEVT nPT(xx.x%) | 0.xxx |
| **SAE ongoing** | nEVT nPT(xx.x%) | nEVT nPT(xx.x%) | 0.xxx |
| **Serious events** |  |  | 0.xxx |
| Death or life-threatening | nEVT nPT(xx.x%) | nEVT nPT(xx.x%) |  |
| Causing or prolonging a hospital stay | nEVT nPT(xx.x%) | nEVT nPT(xx.x%) |  |
| Severe disability or incompetence | nEVT nPT(xx.x%) | nEVT nPT(xx.x%) |  |
| Congenital malformations in offspring | nEVT nPT(xx.x%) | nEVT nPT(xx.x%) |  |
| Important clinical events as judged by the investigators | nEVT nPT(xx.x%) | nEVT nPT(xx.x%) |  |
| **Outcome of the AE** |  |  | 0.xxx |
| Recovered | nEVT nPT(xx.x%) | nEVT nPT(xx.x%) |  |
| Recovered with sequelae | nEVT nPT(xx.x%) | nEVT nPT(xx.x%) |  |
| Death | nEVT nPT(xx.x%) | nEVT nPT(xx.x%) |  |
| **Causal relationship to the treatment received in ED** |  |  | 0.xxx |
| Unrelated (unlikely, not related) | nEVT nPT(xx.x%) | nEVT nPT(xx.x%) |  |
| Related (definitely, probably, possibly) | nEVT nPT(xx.x%) | nEVT nPT(xx.x%) |  |
| **Causal relationship to the study treatment received in ED** |  |  | 0.xxx |
| Unrelated (unlikely, not related) | nEVT nPT(xx.x%) | nEVT nPT(xx.x%) |  |
| Related (definitely, probably, possibly) | nEVT nPT(xx.x%) | nEVT nPT(xx.x%) |  |
| **Expectedness to the study treatment** |  |  | 0.xxx |
| Expected | nEVT nPT(xx.x%) | nEVT nPT(xx.x%) |  |
| Unexpected | nEVT nPT(xx.x%) | nEVT nPT(xx.x%) |  |
| Is there a significant safety issue (SSI) requiring a change to the PIC, protocol, or another urgent safety measure (USM)? | nEVT nPT(xx.x%) | nEVT nPT(xx.x%) |  |
| **Hospitalisation adverse event** |  |  | 0.xxx |
| **Principal diagnosis** |  |  | 0.xxx |
| xxx | xx (xx.x%) | xx (xx.x%) |  |
| xxx | xx (xx.x%) | xx (xx.x%) |  |
| **Hospitalisation related to an SAE** | nEVT nPT(xx.x%) | nEVT nPT(xx.x%) | 0.xxx |
| **Unplanned re-presentation to the ED (7 days to 6 weeks)** | nEVT nPT(xx.x%) | nEVT nPT(xx.x%) | 0.xxx |

Notes:
*p-values obtained from fisher exact test.*

**Table 7:**
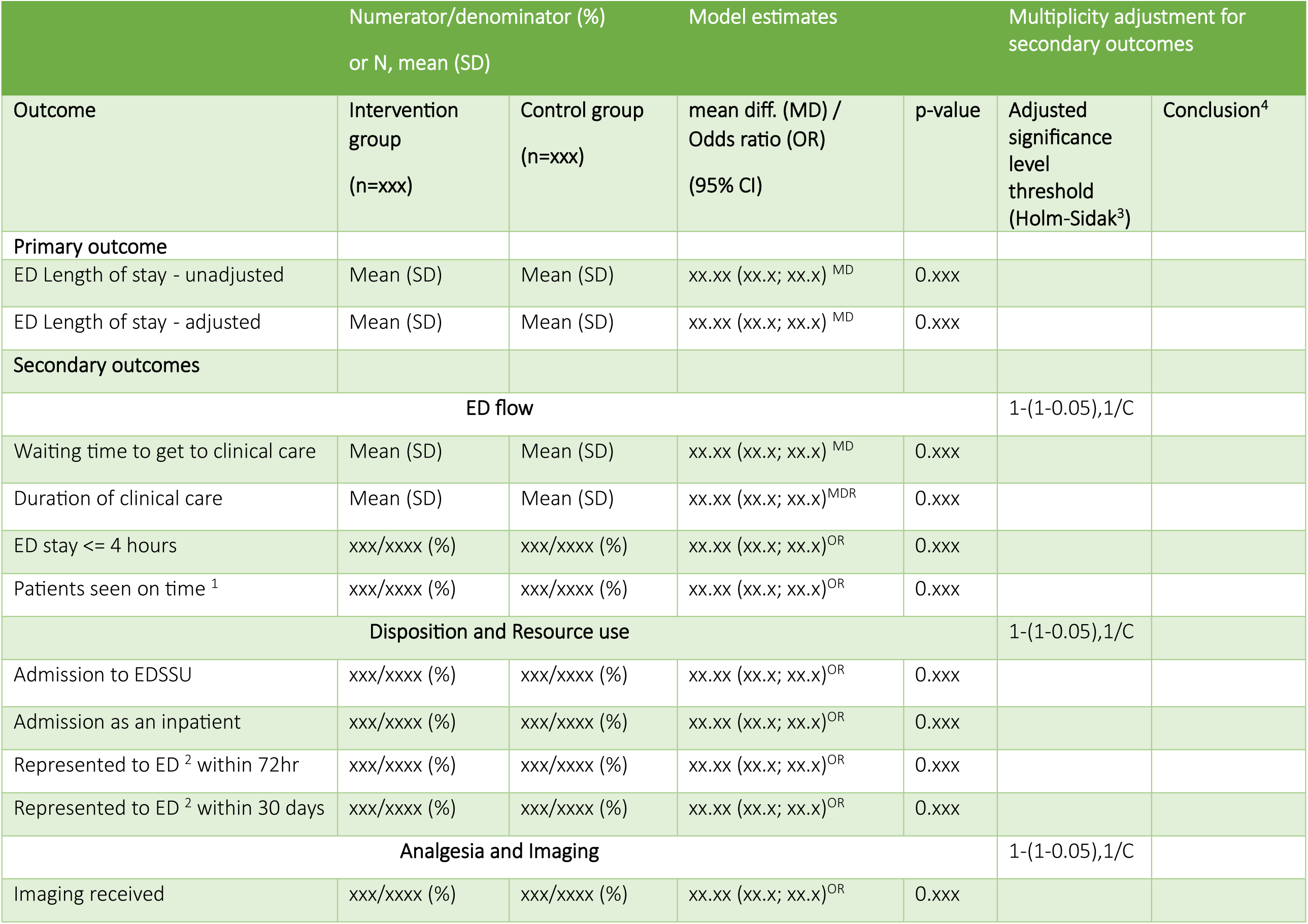
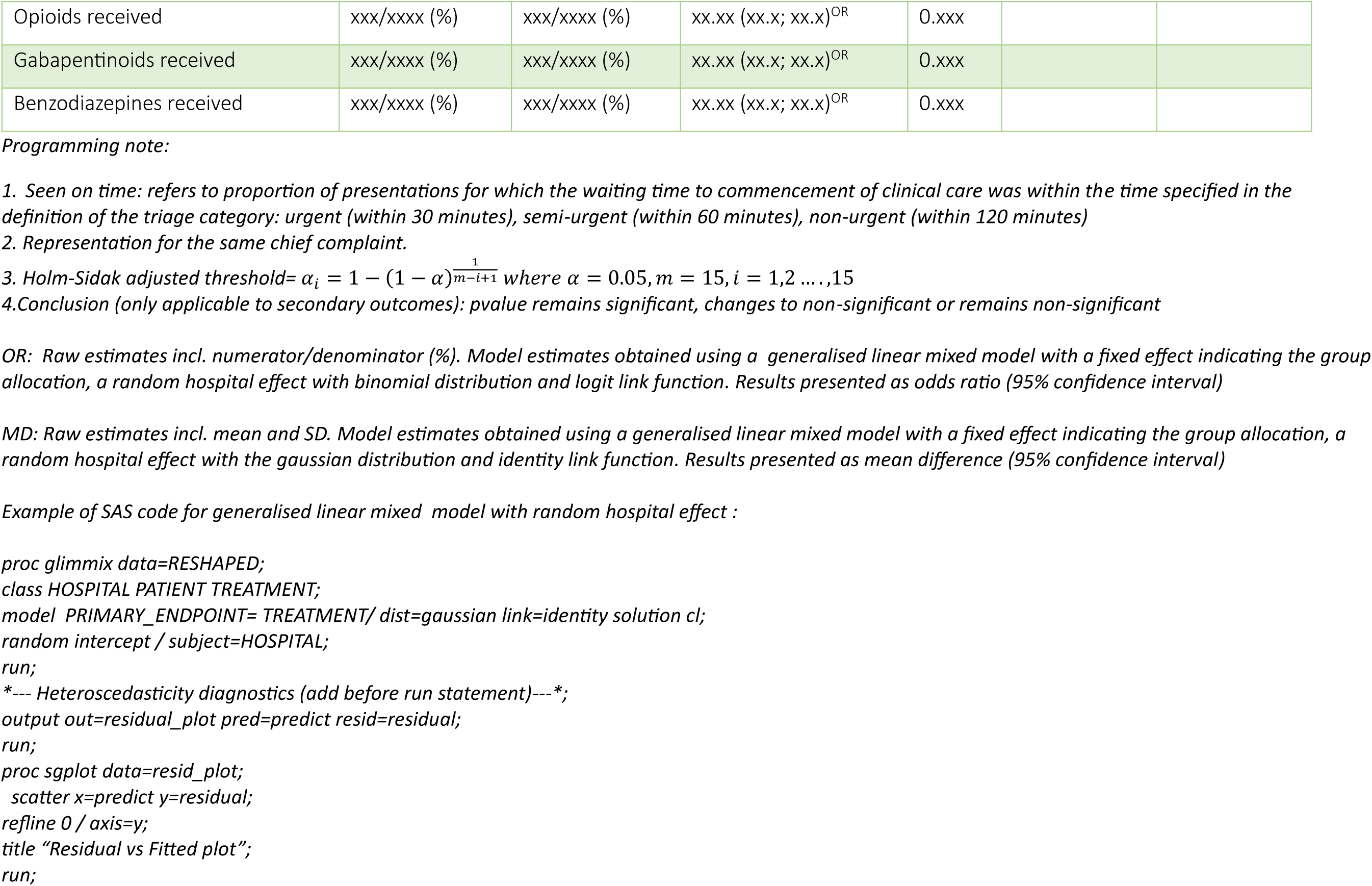
Health services outcomes – model results.

**Table 8:**
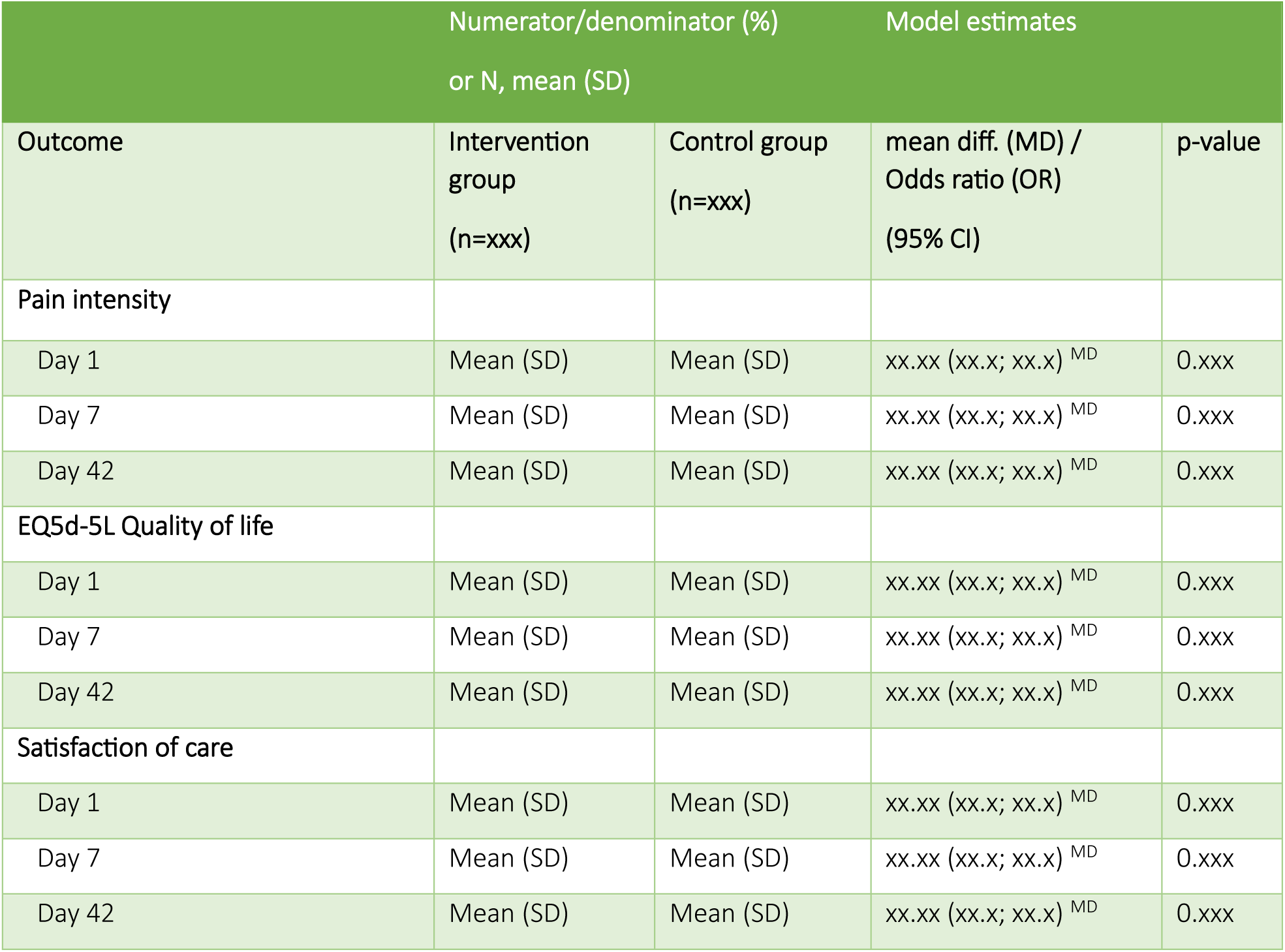
Patient reported outcomes – model results.

**Table 9:**
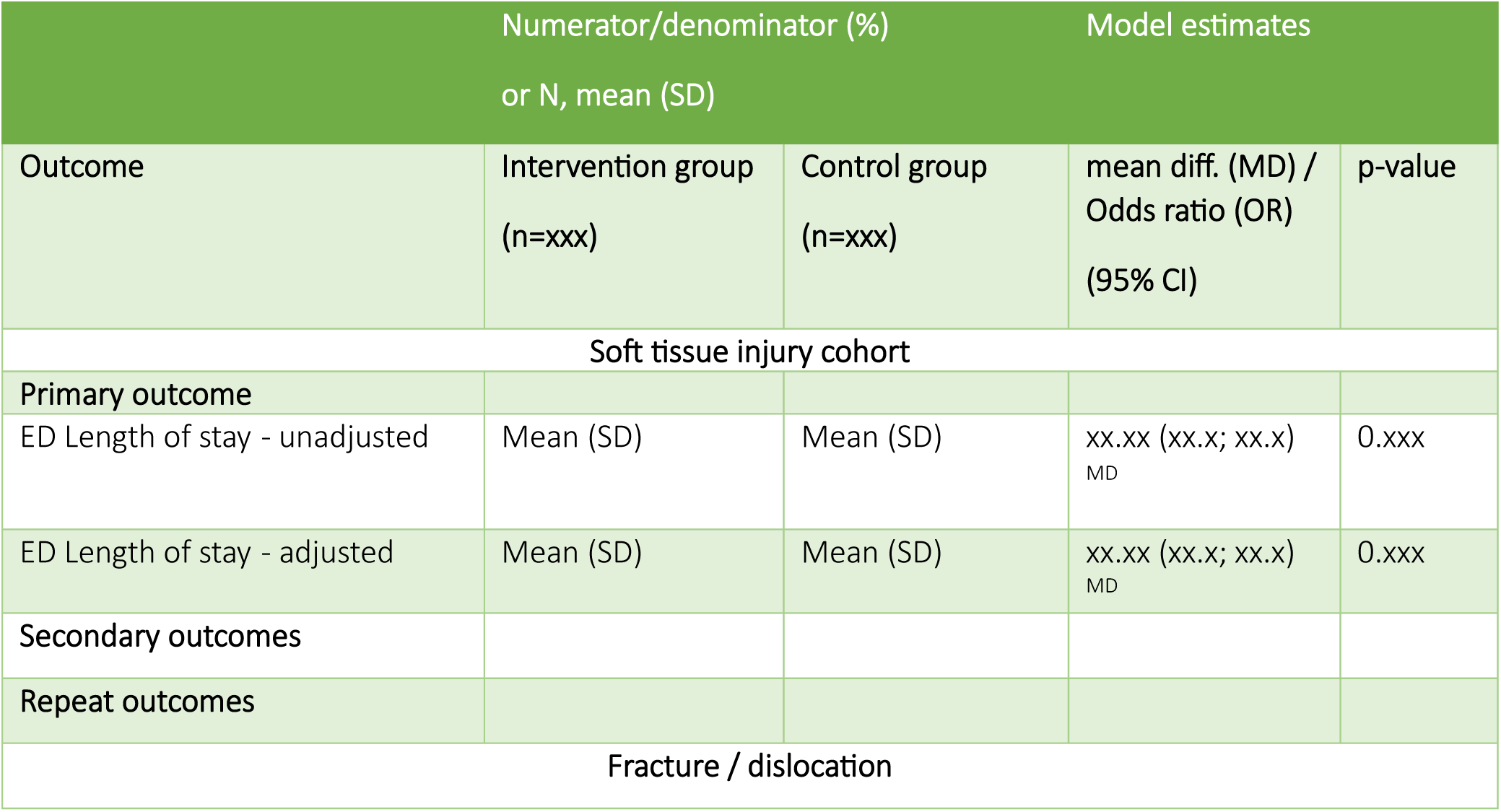
Health services outcomes – model results by MSK condition (repeat Table 7)

**Table 10:**
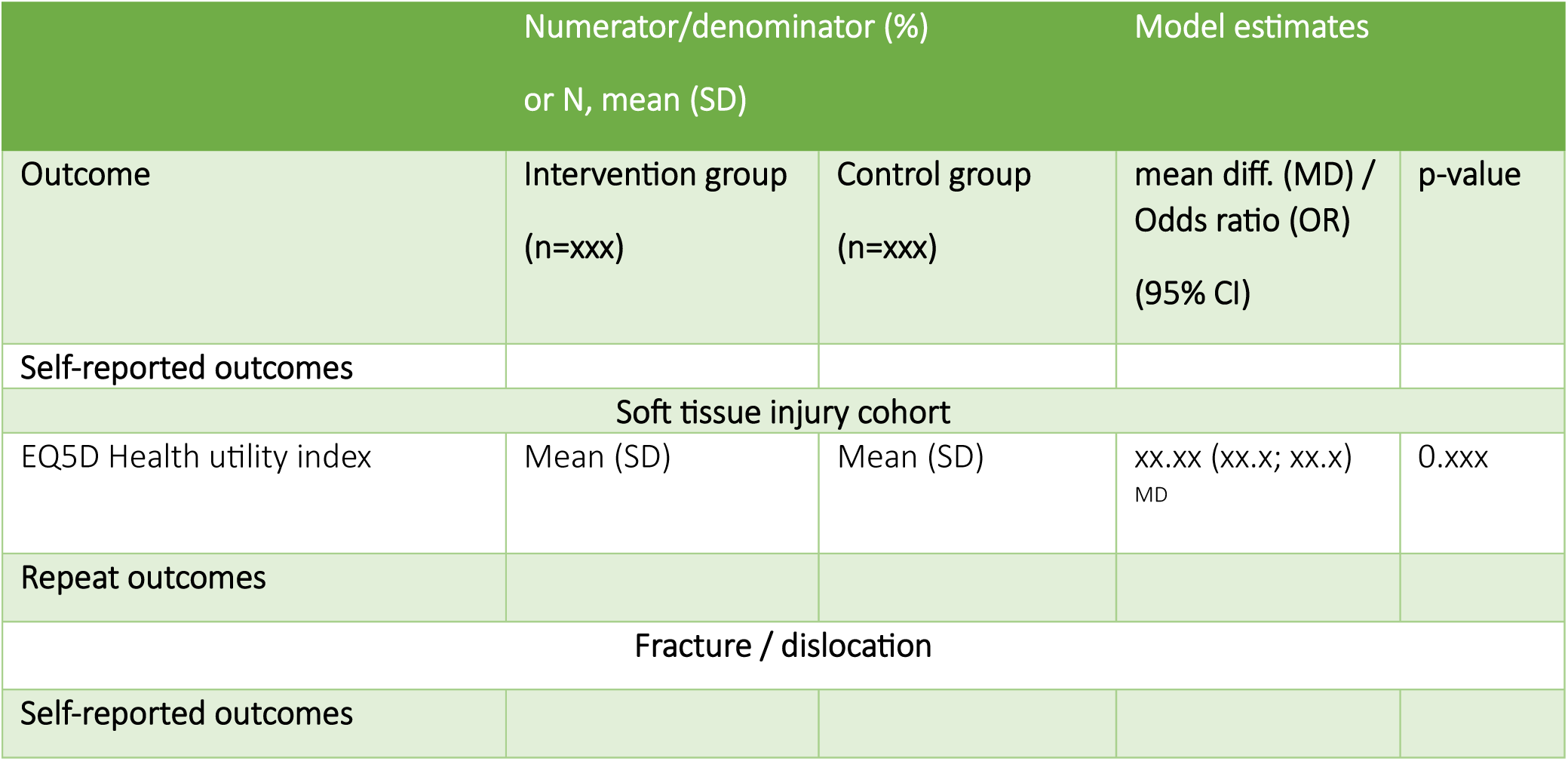
Health services outcomes – model results by musculoskeletal condition (repeat Table 8)

**Listing 1. SAE listing**

**Listing 2: AE listing**

**Listing 3: Protocol deviations**

**Figure 2:** Forest plot for subgroup analysis of primary outcome.

**Figure 3:**
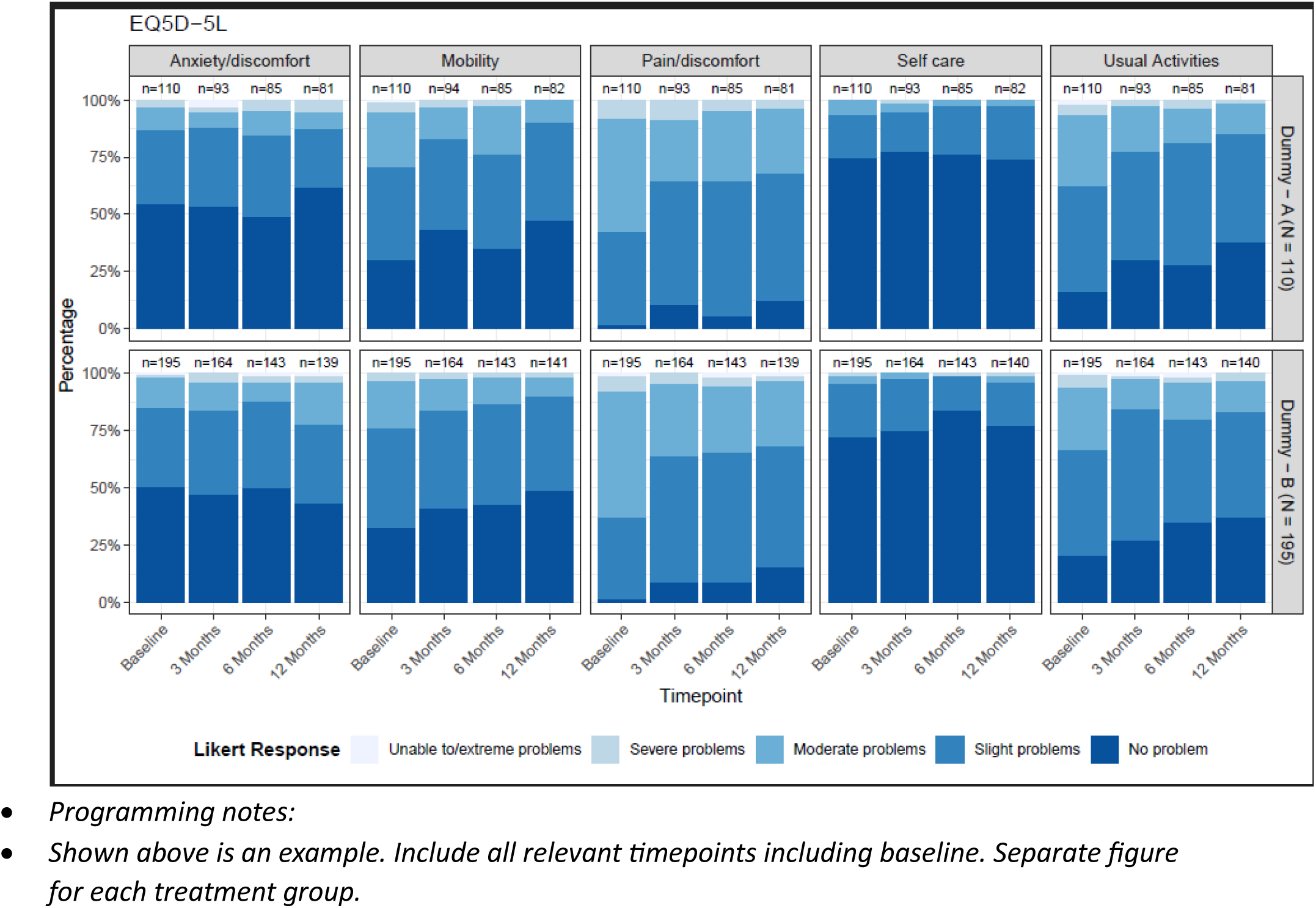
Bar chart of EQ5D-5L across all timepoints (Day 1, Day 7, Day 42)

**Figure 4:** Mean plot of *health utility score (EQ5D)* over time (Day 1, Day 7, Day 42) by treatment groups. *Programming notes:* *Display raw means on the graph as numbers near each dot and denominators below the x-axis*

**Figure 5:** Mean plot of *pain intensity score* over time (Day 1, Day 7, Day 42) by treatment groups.

## Notes

### Competing Interest Statement

The authors have declared no competing interest.

### Clinical Trial

ACTRN 12623000782639; Universal Trial Number (UTN): U1111-1292-2883

### Clinical Protocols

https://pubmed.ncbi.nlm.nih.gov/40074281/

### Funding Statement

The RESHAP-ED trial received funding from the Medical Research Future Fund (MRFF)

### Author Declarations

Sydney Local Health District (RPAH zone) Human Research Ethics Committee (X23-0143).

